# Full-Field Ultrasound Time-Harmonic Elastography Quantifies Task-Dependent Thoracolumbar Fascia and Multifidus Mechanics During Back Extension

**DOI:** 10.64898/2026.09.17.26363289

**Authors:** Eduard Kurz, Tom Meyer, René Schwesig, Karl-Stefan Delank, Ingolf Sack, Hossein S. Aghamiry

## Abstract

**Purpose:** Ultrasound time-harmonic elastography (THE) uses continuous multifrequency vibration and standard-frame-rate ultrasound to generate full-field mechanical maps. We tested whether THE concurrently resolves task-dependent responses in the thin thoracolumbar fascia (TLF) and deeper multifidus muscle (MFM) during back extension.

**Methods:** Eighteen healthy adults (9 female and 9 male; age 19.4 ± 1.9 years) underwent multifrequency THE (60–80 Hz; 100 frames/s) at rest and during active back extension. Shear wave speed (SWS) and penetration rate (PR), an attenuation-sensitive measure, were quantified in manually defined TLF and MFM regions. Four within-tissue contrasts were tested using paired t tests with Holm adjustment; between-tissue, superficial-tissue-thickness, and sex analyses were exploratory.

**Results:** Back extension increased SWS in the TLF (+0.43 ± 0.43 m/s; *dz* = 0.99) and MFM (+0.62 ± 0.23 m/s; *dz* = 2.73) and decreased PR in the TLF (−1.01 ± 0.54 m/s; *dz* = −1.88) and MFM (−1.20 ± 0.51 m/s; *dz* = −2.34); all Holm-adjusted p < 0.001. The SWS increase was 0.19 m/s greater in MFM than TLF (95% confidence interval, 0.01–0.38 m/s; unadjusted p = 0.042), whereas PR changes did not differ between tissues (p = 0.233).

**Conclusion:** THE simultaneously captured spatially resolved, task-dependent responses in adjacent fascial and muscle tissues using standard-frame-rate acquisition. The findings establish biomechanical sensitivity in healthy adults and support further validation with standardized loading, repeatability testing, and comparison with conventional shear wave elastography.

**Clinical trial number:** not applicable.

## Introduction

Low back pain (LBP) is the leading cause of years lived with disability worldwide, affecting an estimated 619 million people in 2020, with the number of prevalent cases projected to increase further by 2050 [1]. Most cases are classified as non-specific, with no single structural cause identifiable by standard imaging [2]. Despite sustained epidemiological and clinical research, the mechanistic underpinnings of the transition from acute to chronic pain remain poorly understood, limiting the development of targeted rehabilitation strategies. Among the paraspinal musculature, the multifidus has received particular attention owing to its role in providing segmental lumbar control. Studies using real-time ultrasound have documented rapid multifidus atrophy following acute LBP episodes, with recovery frequently incomplete even after symptom resolution [3,4]. These structural changes are thought to perpetuate residual dyscontrol and to elevate recurrence risk.

Beyond the contractile elements, the thoracolumbar fascia (TLF) has emerged as a mechanically significant structure in lumbar loading and LBP pathophysiology. The posterior layer of the TLF envelops the erector spinae and multifidus and acts as a tensile force-transmitting interface between the paraspinal muscles, the lumbar vertebrae, and the posterior pelvis [5]. The TLF is densely innervated by nociceptive and sympathetic nerve fibers [6], and experimental hypertonic-saline stimulation of the fascia produces pain of greater magnitude, duration, and radiation than stimulation of the underlying multifidus or subcutaneous tissue [7,8], supporting a role as an independent pain generator rather than a passive envelope. In individuals with chronic LBP, the TLF has been reported to exhibit increased thickness and echogenicity [9], reduced inter-layer shear strain [10], and increased shear stiffness [11] relative to healthy controls, and has been identified as a potential source of pain [12,13]. Beyond its passive tensile role, the fascia may also contribute actively to paraspinal mechanics, since fascial myofibroblasts are capable of smooth-muscle-like contraction [14,15]. These observations suggest that the mechanical properties of the TLF may provide functional information alongside those of the underlying muscle, yet the two structures have not been examined simultaneously within a single quantitative ultrasound elastography examination.

Quantitative ultrasound elastography offers a non-invasive means to characterize soft-tissue viscoelastic properties [16]. A recent systematic review and meta-analysis identified strain imaging, shear wave imaging, and vibration sonoelastography as the principal approaches used to assess back-muscle biomechanics; although reliability was generally good, the authors emphasized limited validity evidence and the need for standardized protocols [17]. Supersonic shear imaging (SSI) has established that shear wave speed (SWS), a surrogate marker of tissue stiffness, increases with isometric contraction in skeletal muscle [18-20]. Complementing SWS, penetration rate (PR) is a frequency-scaled measure inversely related to wave attenuation: lower PR values indicate stronger attenuation and greater dissipative loss [21]. Conventional shear wave elastography commonly uses focused acoustic radiation force impulses and ultrafast acquisition to track transient shear waves. Ultrasound time-harmonic elastography (THE) is a novel full-field method that instead exploits continuous multifrequency mechanical vibrations and encodes steady-state wavefields. Unlike these transient approaches, THE maps spatially and temporally resolved SWS and PR across the full field of view, operates with standard-frame-rate ultrasound rather than requiring an ultrafast scanner, and provides both outputs simultaneously, making it well suited to posture-dependent and prolonged measurements [21-23].

Beyond applications in the brain [24], heart [25], and liver [26], THE has recently been applied to skeletal muscle during isometric exercise, demonstrating rapid full-field stiffness mapping across multiple muscle groups [27]. In the vastus lateralis, THE has further resolved independent contractile and hemodynamic contributions to muscle viscoelasticity during voluntary contraction and blood-flow restriction [28], and has shown that shear-wave anisotropy increases with contraction intensity [29]. That work established orientation-sensitive SWS mapping in a limb muscle under force-controlled contraction. Whether THE can concurrently resolve a thin fascial layer and the deeper paraspinal muscle during trunk loading remains unknown.

Here, we addressed this measurement problem by applying ultrasound THE to the TLF and MFM during back extension. We evaluated whether one multifrequency acquisition could produce co-registered SWS and PR maps in both tissues and detect changes from prone rest to active extension. The four within-tissue condition contrasts constituted the primary analysis; between-tissue, superficial-tissue-thickness, and sex analyses were exploratory. This design links quantitative engineering outputs to biological responses during a standardized trunk-loading task.

## Materials and Methods

### Study Design and Participants

In this pilot study, SWS and PR were compared in the MFM and overlying TLF during prone rest and active back extension. Participants were recruited from among students completing the Federal Volunteer Service at one participating university medical center. Eighteen healthy young adults (9 males and 9 females; Table 1) agreed to participate and provided written informed consent before data collection. Inclusion required no current or recent musculoskeletal injury to the lumbar spine or lower extremities, no history of lumbar surgery, and no episode of LBP in the 12 months preceding the study. Participants refrained from vigorous physical activity for at least 24 h before measurement. The study was approved by the local ethics committees of Charité – University Medicine Berlin (reference number: EA4/040/22) and University Medicine Halle (reference number: 2024-208) in accordance with the Declaration of Helsinki.

**Table 1.** Participant characteristics (N = 18; 9 female and 9 male participants). Values are mean ± SD. BMI, body mass index; SLSF, skin and subcutaneous fat thickness at the probe site

| Group | n | Age (years) | Height (cm) | Mass (kg) | BMI (kg/m<sup>2</sup>) | Body fat (%) | SLSF (mm) |
| --- | --- | --- | --- | --- | --- | --- | --- |
| All participants | 18 | 19.4 ± 1.9 | 177 ± 8.5 | 72.9 ± 11.1 | 23.3 ± 3.0 | 21.6 ± 10.5 | 12.7 ± 3.1 |
| Female | 9 | 20.2 ± 2.0 | 170 ± 5.1 | 68.3 ± 8.8 | 23.6 ± 3.4 | 30.3 ± 6.7 | 13.3 ± 2.6 |
| Male | 9 | 18.6 ± 1.4 | 183 ± 6.0 | 77.5 ± 11.6 | 23.1 ± 2.8 | 12.9 ± 4.5 | 12.1 ± 3.6 |

### Measurement Protocol

Participants lay prone on an examination table with their arms alongside the body (Fig. 1). Two conditions were assessed in fixed order: (1) prone rest, during which participants maintained muscular relaxation; and (2) active back extension, in which participants lifted the upper trunk against gravity without hyperextending the lumbar spine and held the position during the 4 s acquisition. All participants received the same verbal task instructions; contraction intensity was not quantified by force measurement or electromyography. A minimum rest interval of 60 s separated conditions.

**Fig. 1.**
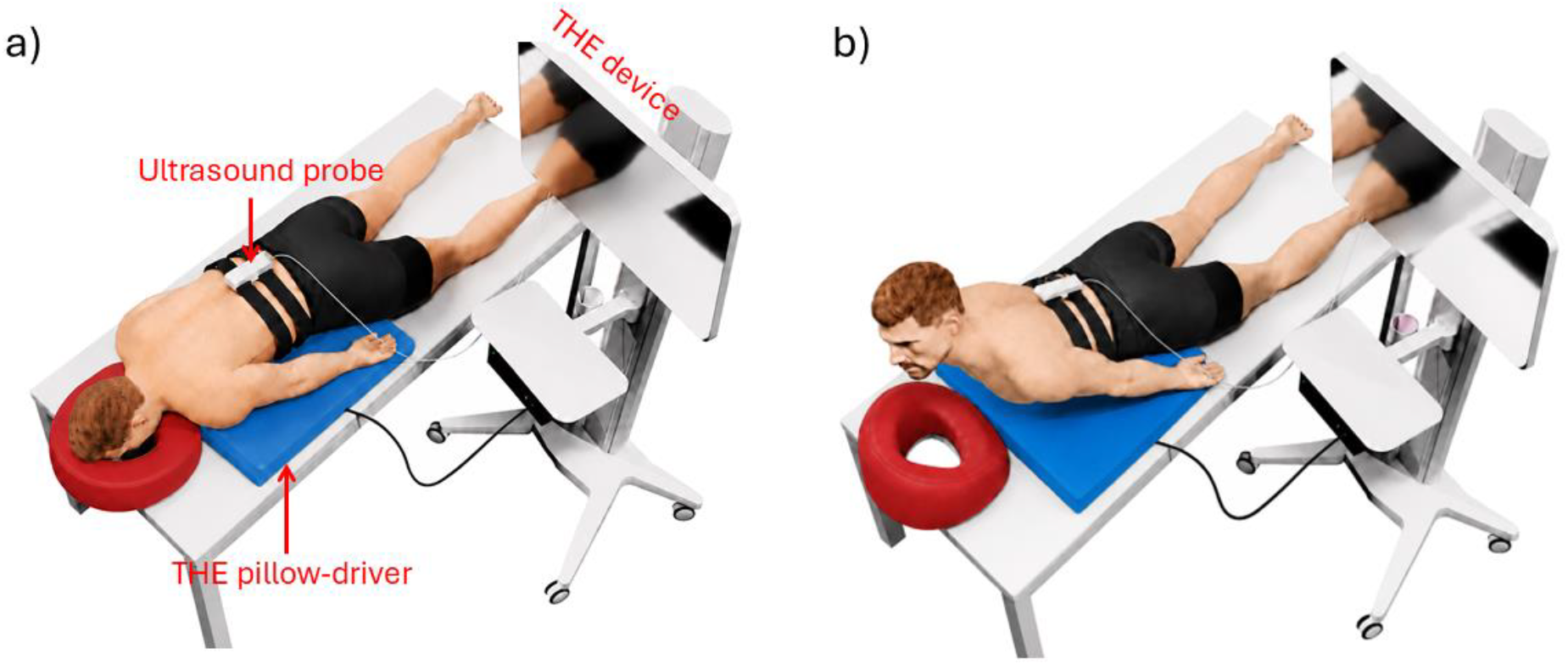
Measurement setup of THE in MFM and TLF. (a) Resting position. (b) Back extension task. The probe holder is secured with elastic bands over the right lumbar paravertebral muscles at L3. The different measurement components are indicated in panel (a)

### THE Measurement System and Acquisition

THE was performed following published protocols [23,28,29]. A custom pillow-driver placed beneath the trunk delivered continuous mechanical vibrations at three superimposed frequencies (60, 70, and 80 Hz). A clinical ultrasound system (GAMPT, Merseburg, Germany) equipped with a linear transducer (5–11 MHz; 54.6 mm footprint) was positioned longitudinally over the right paraspinal musculature at the L3 level (Fig. 1). Radiofrequency data were acquired line-by-line at 100 frames s^−1^ for 4 s per condition, yielding 400 frames per acquisition.

### Signal Processing and Region-of-Interest Definition

Wavenumber-based multifrequency dual elasto-visco (k-MDEV) inversion [21] was applied in five sequential steps: (1) axial tissue displacements were estimated using a monogenic phase-constancy optical-flow solver in a multiscale band-pass pyramid [29]; (2) aliased frequency components arising from the 100 Hz frame rate were realigned to the true excitation frequencies by controlled-aliasing reconstruction; (3) harmonic components at each frequency were isolated with third-order Butterworth band-pass filters (±3 Hz bandwidth); (4) the wavefield at each frequency was decomposed into eight in-plane propagation directions using steerable Gaussian directional filters, suppressing boundary reflections and compressional artifacts; and (5) local wavenumbers were estimated from the spatial phase gradient using k-MDEV inversion. Amplitude-weighted multifrequency averaging yielded SWS maps at approximately 0.25 mm in-plane resolution. PR was defined as PR = ω/k″ (m/s), where k″ is the imaginary part of the complex wavenumber. Pixels with low wavefield quality (z-score |z| > 3 for spectral peak sharpness) were excluded.

Two anatomically distinct regions of interest (ROIs) were delineated manually on the first B-mode frame of each acquisition: (1) the posterior TLF layer, identified as the thin hyperechoic band immediately superficial to the MFM; and (2) the MFM belly, delineated from the TLF to the posterior vertebral lamina and excluding the hypoechoic deep intramuscular tendon. ROI boundaries were verified by an experienced operator. Each ROI was tracked in subsequent frames via vertical cross-correlation with the first frame. Final SWS and PR values were obtained by temporal and spatial averaging across all 400 frames within each ROI.

SLSF thickness was measured on the same B-mode image as the distance from the skin surface to the superficial border of the TLF, serving as an index of the depth of the signal path from the skin to the first ROI.

### Statistical Analysis

Potential sex differences in demographic and THE variables were assessed using two-sided Student’s unpaired t tests and quantified with Cohen’s d. For the main condition analyses, normality of paired differences (extension minus rest) was assessed using Shapiro–Wilk tests; no deviation from normality was detected (all p ≥ 0.084). Two-sided paired t tests evaluated the four condition contrasts, and Cohen’s *dz* (mean paired difference divided by its standard deviation) quantified effect size. The four p values were adjusted using the Holm procedure. Differences in change between ROIs were tested by paired t tests of participant-level change scores. Resting between-ROI differences were assessed using Wilcoxon signed-rank tests. Pearson correlations, with 95% confidence intervals (95% CI) based on Fisher’s z transformation, characterized associations between ROI changes and between SLSF thickness and 12 THE measures (rest, extension, and change for both parameters and both ROIs). Between-ROI, SLSF, and sex analyses were considered exploratory; unadjusted p values are reported, with Holm-adjusted results across the 12 SLSF correlations and 13 THE/SLSF sex comparisons provided to aid interpretation. All tests were two-sided with α = 0.05.

## Results

All 18 participants completed both conditions without adverse events. Two participants (11%; one female and one male) were left-handed. Demographic characteristics are summarized in Table 1. Mean SLSF thickness was 12.7 ± 3.1 mm (range 7.5–19.0 mm) and did not differ between female and male participants (13.3 ± 2.6 vs. 12.1 ± 3.6 mm; t(16) = 0.85, unadjusted p = 0.405, d = −0.40 for male minus female).

Fig. 2 shows SWS and PR maps from one representative participant, superimposed within the selected TLF and MFM ROIs on the corresponding B-mode images. At rest, mean SWS was 2.67 m/s in the TLF and 2.22 m/s in the MFM; during extension, these values increased to 3.72 and 2.95 m/s, respectively. Mean PR decreased from 4.26 to 3.14 m/s in the TLF and from 3.00 to 2.36 m/s in the MFM. The maps also illustrate spatial heterogeneity within both anatomical regions. These values are presented for illustration only; cohort-level effects were evaluated in all 18 participants.

**Fig. 2.**
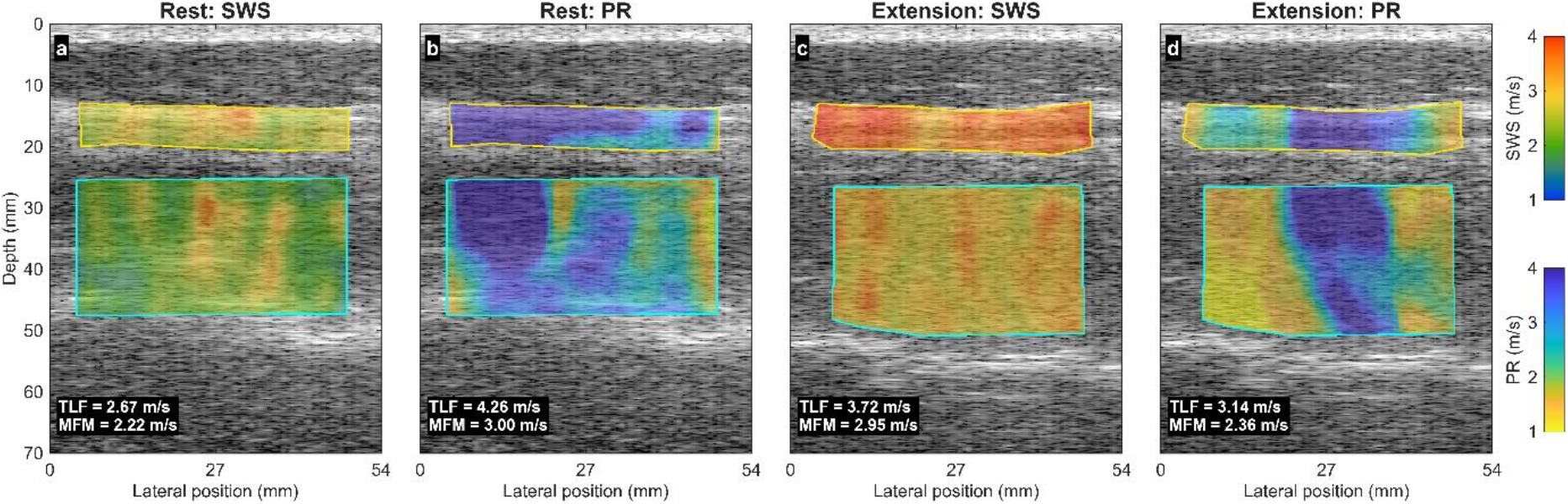
Representative SWS and PR maps superimposed on B-mode ultrasound images from one participant during prone rest and active back extension: (a) SWS at rest, (b) PR at rest, (c) SWS during extension, and (d) PR during extension. Maps are restricted to the selected TLF and MFM ROIs

SWS increased from rest to back extension in both ROIs (Table 2; Figs. 3a and 4a–b). In the TLF, SWS increased from 2.92 ± 0.18 to 3.35 ± 0.36 m/s (ΔSWS = +0.43 ± 0.43 m/s, +14.5%; 95% CI: 0.21 to 0.64 m/s; t(17) = 4.19; *dz* = 0.99). In the MFM, SWS increased from 2.34 ± 0.22 to 2.96 ± 0.33 m/s (ΔSWS = +0.62 ± 0.23 m/s, +26.5%; 95% CI: 0.51 to 0.73 m/s; t(17) = 11.60; *dz* = 2.73). In an exploratory direct comparison, the SWS increase in MFM was 0.19 m/s greater than in TLF (95% CI: 0.01 to 0.38 m/s; t(17) = 2.20, unadjusted p = 0.042; *dz* = 0.52). At rest, TLF SWS was higher than MFM SWS (median paired comparison, Wilcoxon p < 0.001).

**Table 2.** Descriptive statistics and paired condition comparisons for SWS and PR in the TLF and MFM. Δ, extension minus rest; *dz*, paired-sample standardized mean change. Values are mean ± SD. p values are two-sided and Holm-adjusted across the four primary condition contrasts

| ROI | Parameter (m/s) | Rest | Extension | $\Delta$ | Relative change (%) | $d_z$ | Holm-adjusted p |
| --- | --- | --- | --- | --- | --- | --- | --- |
| TLF | SWS | 2.92 ± 0.18 | 3.35 ± 0.36 | +0.43 ± 0.43 | +14.5 | 0.99 | < 0.001 |
| TLF | PR | 4.10 ± 0.45 | 3.09 ± 0.54 | -1.01 ± 0.54 | -24.6 | -1.88 | < 0.001 |
| MFM | SWS | 2.34 ± 0.22 | 2.96 ± 0.33 | +0.62 ± 0.23 | +26.5 | 2.73 | < 0.001 |
| MFM | PR | 3.04 ± 0.59 | 1.84 ± 0.46 | -1.20 ± 0.51 | -39.6 | -2.34 | < 0.001 |

**Fig. 3.**
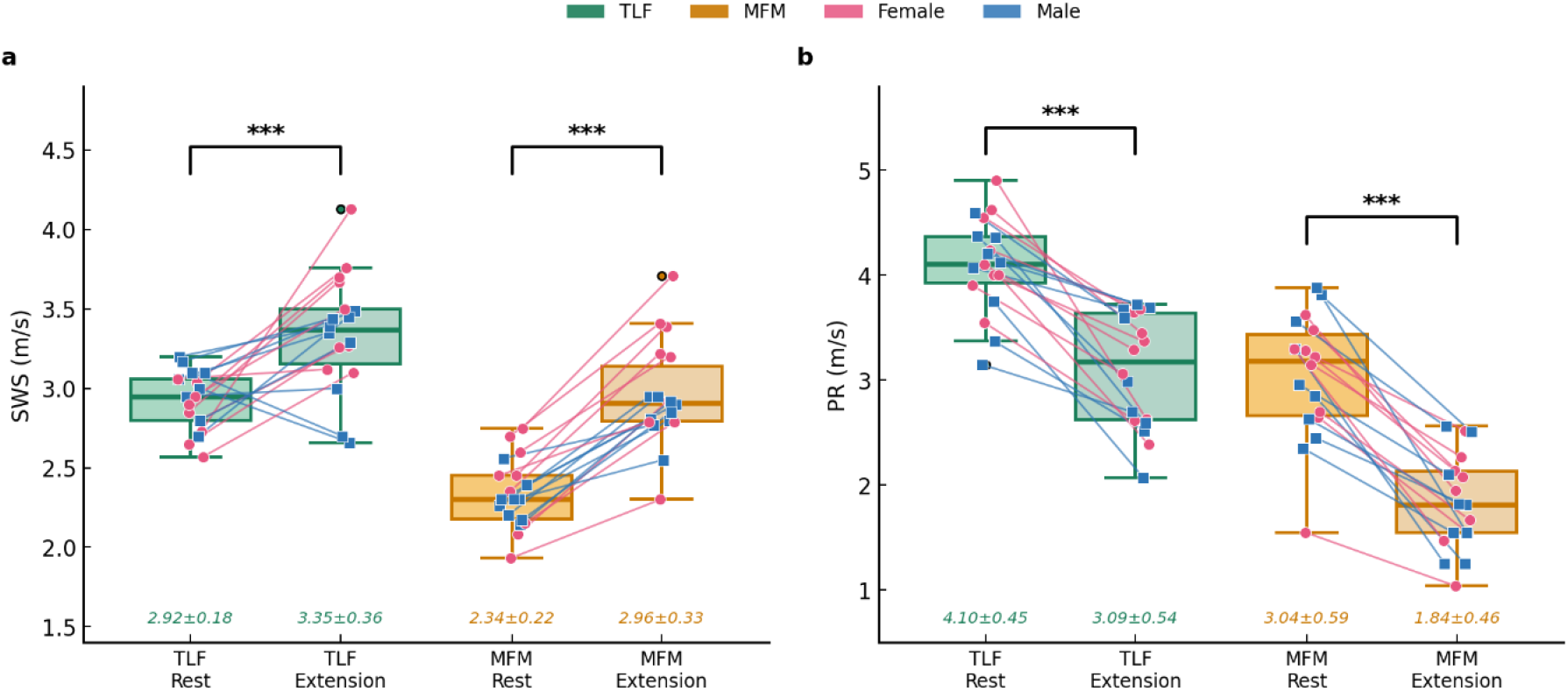
SWS and PR at rest and during back extension: (a) SWS in the TLF and MFM and (b) PR in the TLF and MFM. Box plots show the median, interquartile range, and 1.5× interquartile-range whiskers; individual paired trajectories are coded by sex (pink circles, female; blue squares, male). Annotations report mean ± SD. Significance brackets show the paired t-test results (***p < 0.001)

**Fig. 4.**
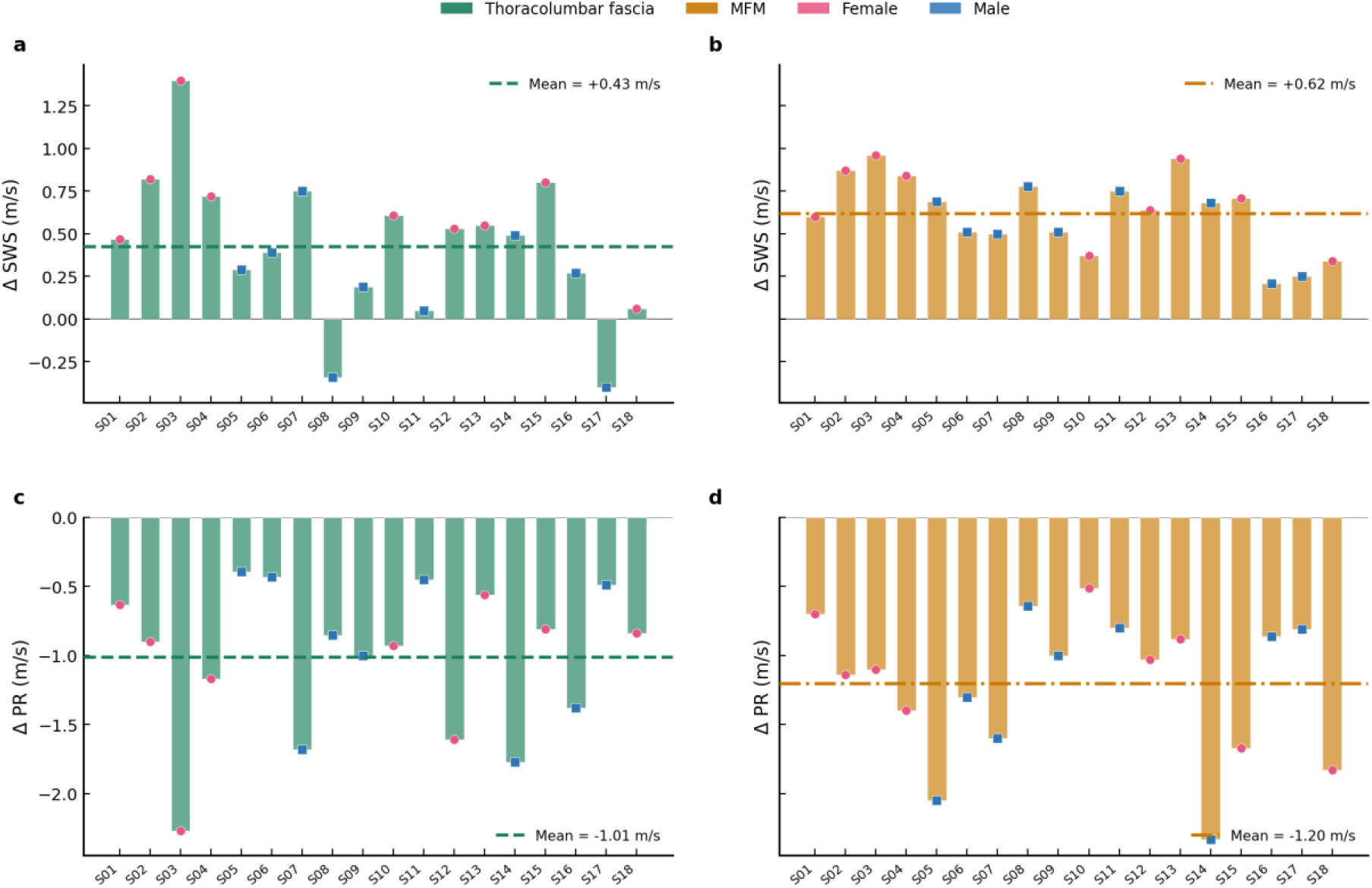
Individual change scores (extension minus rest) for (a) TLF SWS, (b) MFM SWS, (c) TLF PR, and (d) MFM PR across participants S01–S18. Horizontal dashed lines indicate the group mean. Markers are coded by sex (pink circles, female; blue squares, male)

PR decreased during extension in both ROIs (Table 2; Figs. 3b and 4c–d). TLF PR decreased from 4.10 ± 0.45 to 3.09 ± 0.54 m/s (ΔPR = −1.01 ± 0.54 m/s, −24.6%; 95% CI: −1.28 to −0.74 m/s; t(17) = −7.97; *dz* = −1.88). MFM PR decreased from 3.04 ± 0.59 to 1.84 ± 0.46 m/s (ΔPR = −1.20 ± 0.51 m/s, −39.6%; 95% CI: −1.46 to −0.95 m/s; t(17) = −9.94; *dz* = −2.34). Although the relative PR decrease was larger in the MFM than in the TLF, the extension-induced change in PR did not differ significantly between regions (MFM minus TLF: −0.19 m/s; 95% CI: −0.52 to 0.14 m/s; t(17) = −1.24, unadjusted p = 0.233). At rest, TLF PR was higher than MFM PR (Wilcoxon p < 0.001). All four main condition contrasts (i.e. TLF SWS, MFM SWS, TLF PR, and MFM PR) remained significant after Holm correction (adjusted p < 0.001).

The extension-induced SWS changes were positively associated between ROIs (r = 0.488; 95% CI: 0.027 to 0.778; unadjusted p = 0.040; Fig. 5a). The corresponding PR-change association was weaker and imprecise (r = 0.199; 95% CI: −0.295 to 0.609; p = 0.428; Fig. 5b). These exploratory results suggest a possible shared component of the SWS response, but the wide confidence intervals do not establish mechanical coupling or independence between tissue layers.

**Fig. 5.**
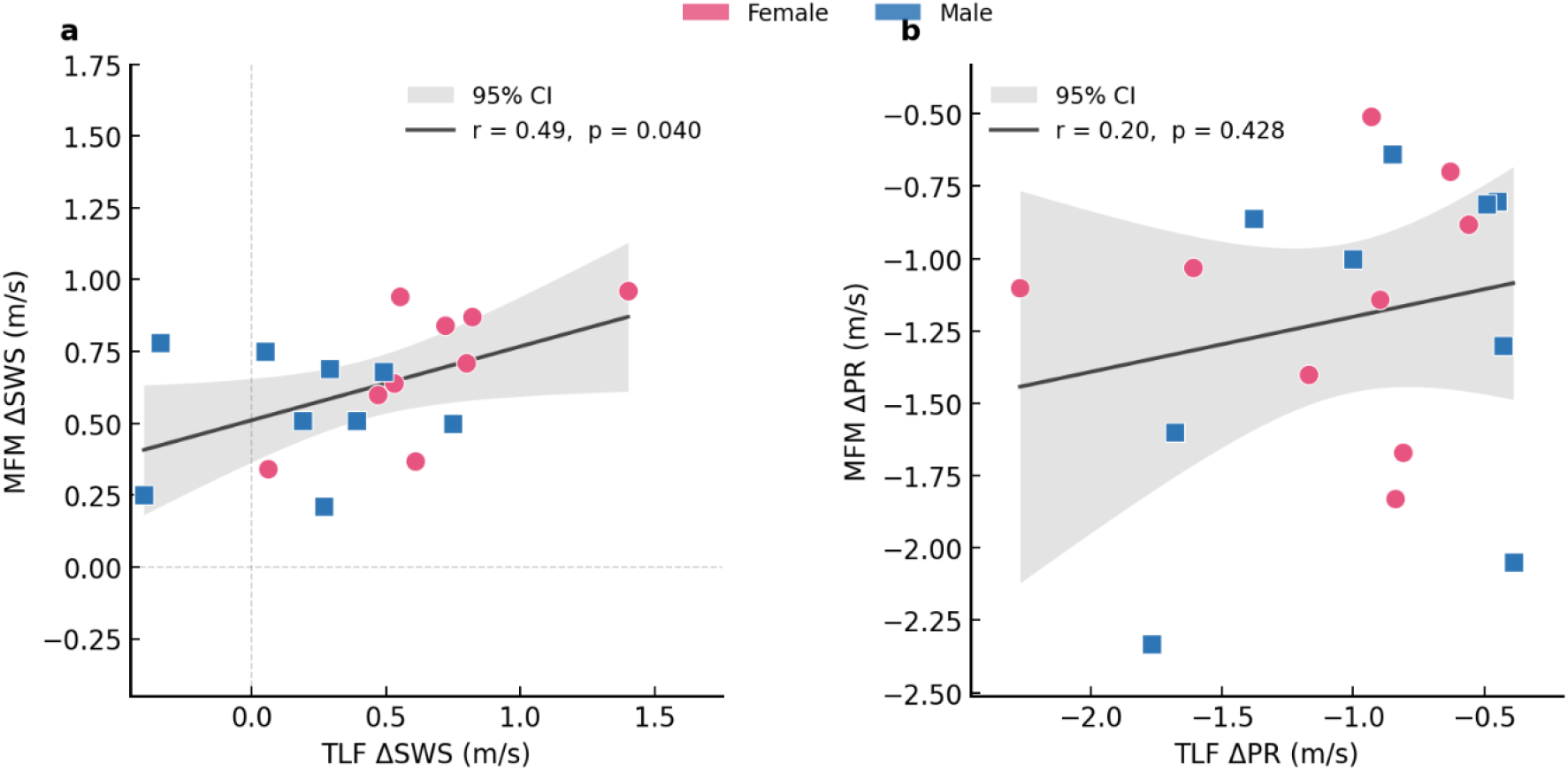
Correlation of extension-induced changes between the TLF and MFM for (a) ΔSWS and (b) ΔPR. Solid lines show linear fits and gray bands show 95% CI for the mean fitted response. Each point represents one participant (pink circles, female; blue squares, male)

SLSF thickness had nominal positive associations with TLF SWS during extension (r = 0.476; 95% CI: 0.011 to 0.771) and with the TLF SWS change (r = 0.495; 95% CI: 0.036 to 0.78; Fig. 6b). However, none of the associations remained significant after Holm correction across the 12 SLSF correlations (adjusted p = 0.507 and 0.442, respectively). TLF SWS at rest was not clearly associated with SLSF thickness (r = −0.222; 95% CI: −0.624 to 0.273; Fig. 6a), and no other SLSF–THE association was strong (|r| ≤ 0.323).

**Fig. 6.**
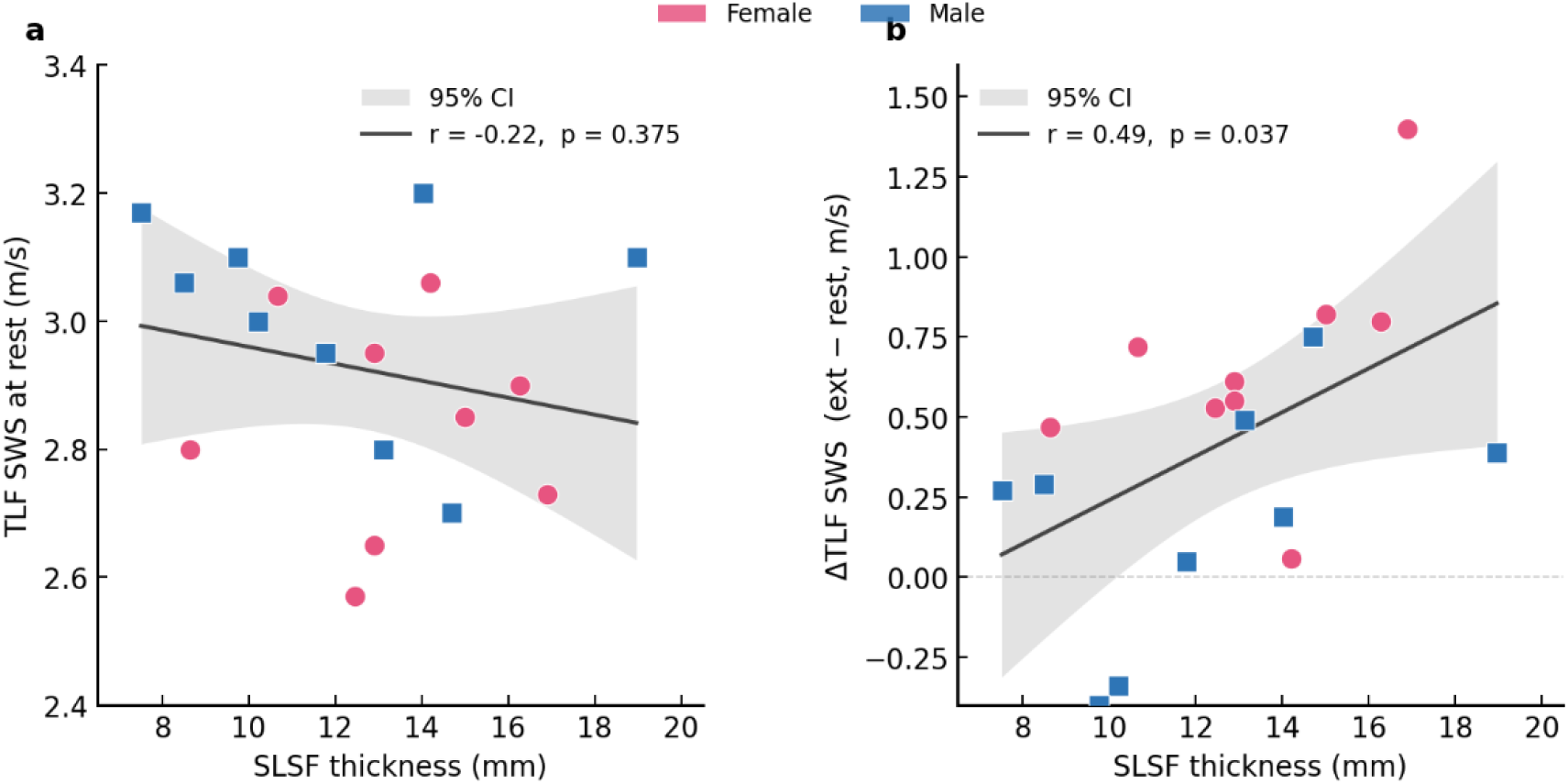
Relationships between SLSF thickness and TLF SWS: (a) SLSF versus resting TLF SWS and (b) SLSF versus the extension-induced TLF SWS change. Solid lines show linear fits and gray bands show 95% confidence intervals for the mean fitted response. Points are coded by sex as in Fig. 5

Exploratory sex comparisons showed higher resting TLF SWS in male than female participants (3.01 ± 0.17 vs. 2.84 ± 0.17 m/s; unadjusted p = 0.047), whereas the TLF SWS increase was larger in female participants (0.66 ± 0.36 vs. 0.19 ± 0.37 m/s; p = 0.014). However, these results were not significant after Holm correction across 13 THE/SLSF sex comparisons (all adjusted p ≥ 0.182) and should therefore not be interpreted as established sex effects.

## Discussion

This proof-of-concept study addressed a biomedical measurement challenge: concurrent quantification of task-dependent mechanics in a thin fascia and an underlying muscle. Full-field ultrasound THE detected increased SWS and decreased PR in both the TLF and MFM during prone low-level back extension, with large within-participant standardized effect sizes. These findings establish sensitivity to mechanical responses across adjacent tissues with different architecture within one examination.

The mean SWS increase was larger in MFM than TLF (+0.62 vs. +0.43 m/s), and the exploratory paired comparison of change scores suggested a modest between-ROI difference (+0.19 m/s; unadjusted p = 0.042). This pattern is compatible with an additional contribution from active actin–myosin cross-bridge formation in the muscle belly, while the TLF is likely loaded through tension in its collagen architecture and coupling to the contracting musculature. However, THE cannot separate active from passive mechanisms. Fascial myofibroblasts can contract in a smooth-muscle-like manner [14,15], but their reported time course is much slower than the acute task used here. The higher resting TLF SWS is consistent with a mechanical difference between dense connective tissue and relaxed muscle, although SWS is also influenced by anisotropy, loading, and acquisition geometry.

PR decreased substantially during extension in both ROIs. The relative reduction was descriptively larger in the MFM (−39.6%) than in the TLF (−24.6%), but the direct difference in participant-level PR changes was not statistically clear (p = 0.233). In the k-MDEV definition used here, PR = ω/k″ is proportional to attenuation length at a given angular frequency; a lower value therefore indicates stronger attenuation, but it should not be interpreted as a literal propagation distance or as a direct measure of viscosity. Possible contributors include fluid–matrix interactions, fiber orientation relative to wave propagation, and intramuscular pressure. Likewise, the nominal ΔSWS correlation between ROIs may reflect a shared loading component, but its wide confidence interval and unadjusted p value do not establish mechanical coupling. The non-significant ΔPR correlation also cannot demonstrate that the two attenuation responses are independent.

Resting MFM SWS of 2.34 m/s corresponds to an apparent shear modulus of approximately 5.7 kPa from G = ρv^2^ if a density of 1050 kg/m^2^ and an isotropic, incompressible, non-dispersive medium are assumed. This is close to conventional shear-wave elastography values widely reported for the lumbar multifidus in healthy adults: 6.1 kPa [20], 5.8 kPa [31], and 5.4 ± 1.6 kPa [32]. The agreement is encouraging but should be interpreted cautiously because THE and SSI use different excitation frequencies, wavefields, inversion methods, and sampling geometries. Muscle is anisotropic, viscoelastic, and dispersive, so formal cross-method validation in the same participants is needed before the values can be treated as interchangeable.

During back extension, MFM SWS rose to 2.96 m/s, corresponding to an apparent modulus of approximately 9.2 kPa under the same simplifying assumptions. This is lower than the approximately 24 kPa reported by Koppenhaver et al. [20] during a standardized contralateral arm lift at about 30% maximal voluntary isometric contraction and the approximately 30 kPa observed by Masaki et al. [31] during trunk flexion. The difference is plausible because the present task was an unresisted postural lift without force normalization, but differences in acquisition and inversion also preclude direct comparison. In unilateral lumbar disc herniation, Alis et al. [33] reported values around 14 kPa on the affected side and 18–19 kPa on the unaffected side in an older clinical cohort. Increased TLF and paraspinal muscle stiffness has also been reported in chronic non-specific LBP [34] and in weightlifters with LBP [11]. These observations support clinical evaluation of the protocol, while PR offers a complementary attenuation-sensitive outcome whose physiological interpretation still requires validation.

SLSF thickness showed nominal positive correlations with extension TLF SWS and with the TLF SWS change. The confidence intervals were wide, and neither result survived correction across the 12 SLSF correlations. The data therefore identify a possible depth- or anatomy-related influence rather than a TLF-specific effect. This possibility is relevant because external vibrations and the ultrasound beam pass through the superficial tissues before reaching the TLF, but it should be tested prospectively in a larger cohort with a broader range of body composition.

From a biomedical engineering perspective, continuous external multifrequency actuation, standard-frame-rate radiofrequency acquisition, and k-MDEV inversion produced co-registered SWS and PR maps from one examination. Concurrent analysis of the thin TLF and deeper MFM shows that the pipeline can resolve task-dependent responses across adjacent tissues with different geometry and composition. The present study evaluates sensitivity rather than measurement accuracy or reliability; phantom comparison, repeated acquisitions, interoperator testing, probe-pressure control, and comparison with conventional shear wave elastography remain necessary for full technical validation.

Although encouraging, our study had limitations. The sample was small and restricted to healthy young adults, limiting the generalizability of our results and providing insufficient power for stable sex-stratified or correlation estimates. Rest always preceded extension, so order, fatigue, or adaptation effects cannot be separated from condition effects. Back-extension effort was not normalized by external force or electromyography, and a single loading level and measurement session were used. Repeatability, between-operator variability, and the possible influence of residual probe pressure require dedicated study. Manual ROI definition and the differing architecture of superficial and deep MFM fibers [30] may also contribute to measurement variability. Finally, the exploratory analyses involved multiple comparisons; nominal p values near 0.05 should be treated as hypothesis-generating rather than confirmatory. Nonetheless, the large within-participant effects provide a strong signal for further validation in standardized and clinical cohorts, extending the demonstrated range of THE applications in skeletal muscle to the back.

In summary, ultrasound THE generated spatially resolved SWS and PR maps and detected clear task-dependent responses in both the TLF and MFM during prone low-level back extension. SWS increased and PR decreased in both tissues, with large paired effects after correction for the four primary comparisons. These results establish the biomechanical sensitivity of THE for lumbar tissue assessment in healthy adults. Standardized contraction intensity, repeatability and interoperator assessment, cross-method validation, and symptomatic comparison groups are required before the method can support performance monitoring or clinical decisions.

## Acknowledgments

This work was supported by the German Federation of Industrial Research Associations (AiF), project KK5611902 BM4 (MUSKEL), and the German Research Foundation (Deutsche Forschungsgemeinschaft; DFG) through project 513752256 (FOR5628), CRC 1340, GRK 2260 BIOQIC, and grant GU 172614-1. The funders had no role in study design, data collection, analysis or interpretation, manuscript preparation, or the decision to submit the manuscript for publication.

## Statements and Declarations

### Competing Interests

The authors have no relevant financial or non-financial interests to disclose.

### Ethics Approval

This study was performed in accordance with the principles of the Declaration of Helsinki and was approved by the ethics committees of Charité – Universitätsmedizin Berlin (EA4/040/22) and University Medicine Halle (2024-208).

### Data Availability

The datasets generated and analyzed during this study are not publicly available because the applicable ethics approvals do not permit public data sharing. Requests may be directed to the corresponding author and will be considered subject to institutional and ethics requirements. Custom THE processing code is available from the corresponding author on reasonable request.

## Notes

### Competing Interest Statement

The authors have declared no competing interest.

### Author Declarations

The study was approved by the local ethics committees of Charite - University Medicine Berlin (reference number: EA4/040/22) and University Medicine Halle (reference number: 2024-208) in accordance with the Declaration of Helsinki.

